# Seasonal Dynamics of Human IgG Antibody Responses to Aedes Nterm-34kDa Peptide as a Biomarker of Alphavirus and Flavivirus Transmission in Northern Tanzania

**DOI:** 10.1101/2025.11.20.25340645

**Authors:** Debora C. Kajeguka, Neema Kulaya, Emmanuel T. Issangya, Basiliana Emidi, Nancy A. Kassam, Emmanuel Elanga-Ndille

## Abstract

**Background:** Continued expansion of arboviral diseases poses a serious threat to public health systems. The use of biomarkers for mosquito exposure, such as antibodies against the Aedes Nterm-34 kDa peptide, serves as a proxy for assessing the risk of Aedes-borne diseases. This study aimed to evaluate whether IgG antibodies against the Nterm-34 kDa peptide are associated with the level of human exposure to Aedes mosquito bites and the risk of flavivirus and alphavirus infections.

**Methods:** Three longitudinal surveys were conducted during the rainy, dry, and short rainy seasons in three villages in Bondo, Tanga. The study included individuals aged between 2 to 70 years. A questionnaire was administered to participants to collect information on socio-demographic factors, housing conditions, surrounding environmental features, and mosquito bite prevention measures. In parallel, intravenous blood was collected from each participant for virus detection using RT-qPCR and for quantification of IgG antibodies against the Aedes Nterm-34kDa peptide using an ELISA assay

**Results:** A total of 362 participants were enrolled, with a mean age of 28.3 ± 23.1 years. Alphavirus positivity peaked in the rainy season 19 (5.2%), with lower prevalence during dry 4 (1.1%) and short rainy 9 (2.5%) seasons, whereas flavivirus positivity was lower overall 11 (3.0%) rainy, 3 (0.8%) dry, 5 (1.4%) short rainy). IgG antibody levels against the Aedes 34 kDa peptide were significantly higher in PCR-positive. The level of IgG antibody response to the Nterm-34kDa peptide exhibited seasonal variation, with significantly higher values observed during the rainy season compared to the dry season (Mann-Whitney U test, *p* < 0.0001). IgG seropositivity was significantly associated with proximity to vegetation (*p* < 0.001), presence of garbage pits (*p* = 0.005), and recent travel history (*p* = 0.018), whereas demographic and housing factors showed no significant associations. Participants positive for dengue demonstrated higher levels of anti-salivary IgG compared with dengue-negative participants (*p* = 0.02; non-parametric Mann–Whitney test).

**Conclusions:** This study reveals distinct seasonal dynamics in human IgG antibody responses to the Aedes Nterm-34kDa salivary peptide, reflecting the seasonal transmission patterns of Alphavirus and Flavivirus in northern Tanzania. Elevated seropositivity during the rainy season aligns with increased vector exposure driven by environmental factors near human dwellings. The findings underscore the potential of the Nterm-34kDa peptide as a biomarker for arboviral transmission monitoring and highlight the importance of targeted vector control strategies considering seasonal and microenvironmental variations to mitigate Aedes-borne viral infections in the region.

## Background

Arboviral diseases, primarily transmitted by *Aedes* mosquitoes, pose a significant health threat in tropical regions. Diseases such as dengue, Zika, and chikungunya are now recognized as emerging threats with epidemic and pandemic potential, endangering public health and resulting in substantial socio-economic burdens (Tajudeen et al., 2021; WHO, 2024a;. WHO, 2019). The potential epidemiological risk is attributed to the recent increase in the prevalence, incidence, complications, and severity of arboviral diseases with an estimated four billion people at risk of infection from arboviruses worldwide. This number is projected to rise to five billion by 2050 (WHO 2024b). Notably, arboviral diseases cause a serious broad spectrum of manifestation such as haemorrhagic fever, shock syndrome, chronic joint pain, microcephaly and multi-system failure (Chauhan et al. 2022; Milhim et al. 2022).

Aedes-borne diseases continued expansion beyond their traditional geographic boundaries is driven by factors such as urbanization, globalization, climate change, and increased international travel (Subhadra et al. 2021; WHO 2024b; Wilder-Smith et al. 2017). The co-circulation of multiple arboviruses in regions like Africa, Asia, and the Americas has led to severe epidemics, exacerbating the burden on already strained healthcare systems (WHO 2023a). Furthermore, the lack of effective vaccines or treatments for many of these diseases underscores the urgent need for robust surveillance systems, vector control strategies, and investment in research to mitigate their impact (Weaver 2013; WHO 2024b). As arboviral diseases continue to cross borders and affect naïve populations lacking immunity, they pose a persistent threat to global health security. Africa has been a significant epicenter for arboviral diseases, with dengue being one of the most prevalent. Recent data highlights the increasing on-going transmission and severity of dengue outbreaks in Africa, with countries such as Burkina Faso, Côte d’Ivoire, Ethiopia, Kenya, Benin, Mali, and Senegal experiencing major epidemics (WHO 2014, 2023b). Tanzania, as many African countries has experienced repeated outbreaks of dengue in 2010, 2013, 2014, 2018 and 2019 (WHO 2019). There is evidence of active circulation of chikungunya, dengue, zika, rift valley fever and yellow fever viral genomes in the mosquito and human populations (Boillat-Blanco et al. 2018; Kaaya et al. 2023; Kajeguka et al. 2023; Mboera et al. 2016; Philbert and Msonga 2020). *Aedes aegypti* is the primary vector responsible for transmitting dengue and other arboviruses in Tanzania (Philbert and Msonga 2020).

Despite the fact that most of these are neglected diseases, they are preventable through effective control of mosquito vectors. This control can be achieved through management of breeding sites, indoor residual spraying and to conduct extensive surveillance of these mosquito populations to estimate risk of transmission (Achee et al. 2019; Poinsignon et al. 2025a). Assessment of the vector control programs against dengue, zika and chikungunya, is based on entomological methods, that can broadly be categorized into 1) immature stage (larvae and pupae) survey indices, 2) eggs per ovitrap per week, 3) female mosquitoes per sticky gravid trap per week, and 4) adult infection rates (Barbazan et al. 2008; CDC 2017). The most commonly used indicators for evaluating the abundance of *Aedes* population includes the indices of Breteau, Adult Productivity, and Adult density (Focks 2003). The public health importance of these indicators is based on the hypothesis that greater abundance of *Aedes* mosquitoes increases the risk of transmission; hence, reducing human-vector contact decreases the incidence of infection (Cromwell et al. 2017). However, these indicators have several limitations in the larger scale applications. The count of positive breeding sites is labor intensive and time consuming to obtain solid results and the evaluation of adult *Aedes* density by usual entomological methods is not sensitive enough to estimate the low-level exposure to vector bites (Sagna et al. 2018). Estimation of adult *Aedes* mosquitoes’ abundance is mainly appropriate to assess transmission risk, however adults collection related to human lading catches may rise ethical concern (Elanga-Ndille et al. 2016). To increase the efficacy of vector control programs, an epidemiological methodology has been documented (Sagna et al. 2018). This methodology can reliably measure arboviral transmission dynamics that enables more accurate and useful predictions of outbreaks. This method can be used to identify areas of risk for *Aedes*-borne diseases and evaluate vector control interventions (Elanga-Ndille et al. 2014, 2016; Kassam et al. 2022; Olajiga et al. 2022; Sagna et al. 2018). The documented innovative tool is based on measuring human IgG responses to salivary proteins of *Aedes* vectors injected during blood meal. This methodology can reliably measure arboviral transmission dynamics that enable more accurate and useful predictions of outbreaks. Quantification of the human Ab response to mosquito salivary proteins are important correlates with mosquito density or ongoing biting activity. This method can be used to identify areas of risk for *Aedes*-borne diseases and evaluate vector control interventions.

Despite the progress made in understanding the role of *Aedes* salivary biomarkers in assessing human exposure to mosquito bites and immune responses several gaps remain (Doucoure et al. 2014; Elanga-Ndille et al. 2016; Fustec et al. 2021; Kassam et al. 2022; Londoño-Rentería et al. 2015; Mathieu-Daudé et al. 2018). There is limited research directly linking these biomarkers to actual infection risk or arboviral disease outcomes in diverse populations (Londono-Renteria et al. 2013). While several reports have recommended the use of salivary biomarkers, emphasizing their potential applications in evaluating arboviral transmission risk, empirical data demonstrating their utility in real-world settings is still lacking (Etienne et al. 2023; Sagna et al. 2018). Therefore, the present study assessed whether individuals exposed to *Aedes* mosquitoes presents specific anti-IgG antibodies against *Aedes* salivary peptide that could be useful in assessing the risk of alphavirus and flavivirus transmission in northern-eastern Tanzania.

## Material and Methods

### Study area and design

A cross-sectional study was conducted in Bondo area, Tanga, in northeastern Tanzania). Tanga experiences a tropical climate characterized by significant annual rainfall, averaging over 1,212 mm. The region has distinct rainy seasons, with the long rains occurring from March to May, peaking in April and May with over 470 mm of precipitation, and the short rains from October to November, which typically yield more than 250 mm (NDF 2014). Tanga is well-known for its history of recurring dengue and chikungunya outbreaks, highlighting its status as a hotspot for these arboviral infections (Abas et al. 2024; Kajeguka et al. 2016; Philbert and Msonga 2020). The area is characterized by a widespread presence of Aedes aegypti and Aedes albopictus mosquitoes, particularly in and around the seaport area, with vector populations reaching their highest levels during the rainy seasons (Abas et al. 2024). The region’s ecological conditions and epidemiological patterns, marked by high vector densities and frequent arboviral occurrences, make it a potential area for study (Abas et al. 2024; Kajeguka et al. 2016; Philbert and Msonga 2020). Three villages—Bondo, Kwamgwe, and Kwadoya—were chosen, each exhibiting different levels of exposure to Aedes vectors.

### Sample size calculation

The sample size for this study was calculated using the following formula n = (Z^2^ * p * (1-p) / d^2^. In this equation, *n* is the sample size, *z* is the value of the standard normal distribution at the 5% level (1.96), *p* is the prevalence, *q* = 1 – *p*, and *d* is the precision level. The prevalence of arboviral diseases in Tanzania is 38.2% (Ward et al. 2017). The sample size for human exposure to *Aedes* mosquito (as measured by level of anti-*Aedes* salivary proteins antibodies) was 362.

### Participant Recruitment and Interview

Participants from each village were invited to participate in interviews held at local schools or dispensaries. Data were collected using structured interviews, where the questionnaire was employed. The questionnaire captured various aspects, including demographic information, housing characteristics, travel history, mosquito prevention practices, and the presence of vegetation or water bodies around houses.

### Blood sample collection and assessment of Human exposure to *Aedes* mosquitoes

Approximately 0.4 ml of blood samples were drawn from the participants. The blood samples were placed on Whatman filter paper and properly labelled for analysis by enzyme-linked immunosorbent assay (ELISA). To assess the level of exposure, the salivary peptide Nterm-34 kDa (Genepep SA, St-Jean de Vedas, France) was used as an *Aedes*-specific biomarker to quantify the immune response to *Ae. aegypti* mosquito bites through immunoassays, as previously explained elsewhere (Elanga-Ndille et al. 2012, 2014, 2016; Sagna et al. 2018). Briefly, samples were eluted by incubating in 350 µl of phosphate-buffered saline (PBS) with 0.1% Tween (Sigma-Aldrich, St. Louis, MO) at 4°C for 24 hours. A N-term 34kDa antigen solution (10 µg/mL in 100 µl of PBS) was then coated onto 64 wells of a 96-well Maxisorp plate (Nunc, Roskilde, Denmark) at 37°C for 150 minutes. The plates were blocked with 300 µl of Protein-Free Blocking Buffer (Pierce, Thermo Scientific, France) for 45 minutes at 37°C. Samples were diluted at 1/40 in PBS-Tween 1% and incubated overnight at 4°C in two wells containing peptide and one "no antigen" well (100 µl per well). A mouse biotinylated antibody against human IgG (BD Biosciences, San Diego, CA) was added at a dilution of 1/2000 in PBS-Tween 1% and incubated for 90 minutes at 37°C. Following this, peroxidase-conjugated streptavidin (GE Healthcare, Orsay, France) at a dilution of 1/2000 in PBS-Tween 1% and incubated for an additional 60 minutes at 37°C. Colorimetric development was performed using 2,2′-azino-bis (3-ethylbenzthiazoline-6-sulfonic acid) diammonium (ABTS; Thermo Scientific, France) in a 50 mM citrate buffer (pH=4) containing 0.003% H2O2, and absorbance was measured at 405 nm. Individual results were expressed as the ΔOD value calculated using the formula ΔOD = ODx − ODn, where ODx represents the mean OD values from antigen wells and ODn denotes the OD value from the "no antigen" well. A subject was classified as an “immune responder” if their ΔOD exceeded the cut-off calculated as mean (ΔDOunexposed) + 2SD.

### RNA Extraction and Real-Time One-Step RT-PCR Detection of Flavivirus and Alphavirus

Viral RNA was isolated from human whole blood following the protocol provided by Qiagen RNA Blood Mini Kits (Qiagen, Hilton, Germany). In brief, 30 µL of blood was first lysed and homogenized using a QIAshredder spin column. The lysate was then transferred to a spin column where RNA selectively bound to the QIAamp membrane. Purified RNA was finally eluted with 30 to 100 µL of RNase-free water and stored at −20°C until further PCR analysis. The Aridia Zika/Dengue/CHIK real-time PCR test, in a one-step format, was used to detect the Zika, Dengue and CHIK infection (CTK Biotech, Inc., Poway, California, USA). The lyophilized positive control was rehydrated in 100 *μ*L of supplied PCR-grade water. A total of 5 *μ*L of each extracted RNA, negative control, and positive control were pipetted in the respective wells in the plate. The amplification reaction was performed in an AriaMx Agilent PCR machine (Agilent, Santa Clara, California, USA). The thermocycler conditions started with reverse transcription at 45°C for 15 minutes and an initial denaturation at 95°C for 2 minutes, followed by 45 cycles of denaturation at 95°C for 10 seconds and annealing/extension at 60°C for 50 seconds. Fluorogenic data analysis of the samples and controls was performed by the real-time PCR thermocycler software, according to the manufactureŕs instructions.

### Ethics

The study was approved by the Kilimanjaro Christian Medical University College Research and Ethics Review Committee (CRERC) with certificate number 2492 and the National Institute of Medical Research with certificate number NIMR/HQ/R.8a/Vol.IX/3651. Permission to conduct this study was granted by the district medical officer (DMO) of the Handeni district and the village leaders. Written informed consent was obtained from all participants. Assent involved participants in the age range of 2 to 17 years, who were legally not able to consent by themselves. Parents or guardians consented on behalf of participants who were under 18 years. For participants who were unable to read or write, an impartial witness signed on their behalf. Strict confidentiality was maintained throughout the study, with participant information accessible only to the research team. The study was conducted in accordance with Good Clinical and Laboratory Practices to ensure the highest standards of research integrity and participant safety.

### Statistical analysis

All statistical tests were computed using Statistical Package for Sociol Sciences (SPSS) software and Graph Pad Prism Software (San Diego, CA USA). Pearson’s Chi-squared test was used to compare differences in categorical variables. The non-parametric Kruskal–Wallis test was used for comparison of more than two groups. The Mann–Whitney test was used for comparison of Ab levels of two independent groups and the

## Results

### Socio-demographic Characteristics of the study population

A total of 362 participants were enrolled in the study, with a mean age of 28.34±23.07years. The majority, 187 (51.7%), were aged 18 years and above, and most were female, 209 (57.7%). Overall, 333(92.0%), reported earning less than TZS 200,000 per month, and 325 (89.8%) were farmers. Most participants had primary education, 214 (59.1%). About one-third, 131 (36.2%), were married. The majority of households had up to two people, 211 (58.3%), and most participants, 326 (90.1%), had no travel history. Regarding environmental characteristics, 217 (59.9%) had a garbage pit within 200 metres, and 189 (52.2%) lived near vegetation (within 200 meters). A higher proportion, 209 (57.7%), reported no stagnant water or wells within 200 metres. Socioeconomic assessment showed that 290 (80.1%) were in the low socioeconomic status category. Most houses were built with mud walls, 210 (58.0%), and had iron sheet roofing, 269 (74.3%). The majority of households, 297 (82.0%), had less than five livestock. Bed net utilization was 73% (266) in the rainy season, 74.8% (265) in the dry season, and 73.4% (238) in the short rainy season, (Table 1).

**Table 1:** Factors associated with IgG seropositivity (Responders Non-responder)

| Variable | n (%) | Rainy season (n=362) |  |  | Dry season (n=326) |  |  | Short rainy season (n=328) |  |  |
| --- | --- | --- | --- | --- | --- | --- | --- | --- | --- | --- |
|  |  | Responder | Non-Responder | P-value | Responder | Non-Responder | P-value | Responder | Non-Responder | P-value |
| **Age** |  |  |  |  |  |  |  |  |  |  |
| <5 years | 46 (12.7) | 15 (32.6) | 31 (67.4) | 0.46 | 6 (16.2) | 31 (83.8) | 0.08 | 9 (21.4) | 33 (78.6) | 0.49 |
| 6-17 years | 129 (35.6) | 50 (38.8) | 79 (61.2) |  | 29 (24.4) | 90 (75.6) |  | 16 (13.8) | 100 (86.2) |  |
| 18 and above | 187 (51.7) | 79 (42.2) | 108 (57.8) |  | 24 (14.1) | 146 (85.9) |  | 29 (17.1) | 141 (82.9) |  |
| **Sex** |  |  |  |  |  |  |  |  |  |  |
| Female | 209 (57.7) | 84 (40.2) | 125 (59.8) | 0.85 | 39 (20.4) | 152 (79.6) | 0.19 | 30 (15.2) | 167 (84.8) | 0.46 |
| Male | 153 (42.3) | 60 (39.2) | 93 (60.8) |  | 20 (14.8) | 115 (85.2) |  | 24 (18.3) | 107 (81.7) |  |
| **Income** |  |  |  |  |  |  |  |  |  |  |
| < TZs 200,000/= | 333 (92.0) | 133 (39.9) | 200 (60.1) | 0.83 | 55 (18.4) | 244 (81.6) | 0.64 | 49 (16.3) | 252 (83.7) | 0.76 |
| TZs 200,000 to 1M | 29 (8.0) | 11 (37.9) | 18 (62.1) |  | 4 (14.8) | 23 (85.2) |  | 5 (18.5) | 22 (81.5) |  |
| **Occupation** |  |  |  |  |  |  |  |  |  |  |
| Farmer | 325 (89.8) | 130 (40.0) | 195 (60.0) | 0.78 | 54 (18.6) | 237 (81.4) | 0.23 | 46 (15.8) | 246 (84.2) | 0.61 |
| Employed | 13 (3.6) | 4 (30.8) | 9 (69.2) |  | 0 (0.0) | 12 (100) |  | 3 (23.1) | 10 (76.9) |  |
| Business | 24 (6.6) | 10 (41.7) | 14 (58.3) |  | 5 (21.7) | 18 (78.3) |  | 5 (21.7) | 18 (78.3) |  |
| **Education** |  |  |  |  |  |  |  |  |  |  |
| Primary | 214 (59.1) | 85 (39.7) | 129 (60.3) | 0.35 | 34 (17.4) | 161 (82.6) | 0.82 | 32 (16.4) | 163 (83.6) | 0.66 |
| Secondary | 17 (4.7) | 9 (52.9) | 8 (47.1) |  | 3 (18.8) | 13 (81.3) |  | 1 (7.1) | 13 (92.9) |  |
| Tertiary | 3 (0.8) | 0 (0.0) | 3 (100) |  | 0 (0.0) | 3 (100) |  | 1 (33.3) | 2 (66.7) |  |
| At school/Kindergarten/<br>didn't attend | 128 (35.4) | 50 (39.1) | 78 (60.9) |  | 22 (19.6) | 90 (80.4) |  | 20 (17.2) | 96 (82.8) |  |
| **Marital status** |  |  |  |  |  |  |  |  |  |  |
| Married | 131 (36.2) | 52 (39.7) | 79 (60.3) | 0.78 | 23 (19.0) | 98 (81.0) | 0.95 | 25 (21.2) | 93 (78.8) | 0.09 |
| Divorced | 24 (6.6) | 9 (37.5) | 15 (62.5) |  | 4 (21.1) | 15 (78.9) |  | 3 (14.3) | 18 (85.7) |  |
| Single | 121 (33.4) | 52 (43.0) | 69 (57.0) |  | 19 (17.6) | 89 (82.4) |  | 20 (17.9) | 92 (82.1) |  |
| Other(Cohabiting/child) | 86 (23.8) | 31 (36.0) | 55 (64.0) |  | 13 (16.7) | 65 (83.3) |  | 6 (7.8) | 71 (92.2) |  |
| **Average people in the house** |  |  |  |  |  |  |  |  |  |  |
| Up to 2 people | 211 (58.3) | 88 (41.7) | 123 (58.3) | 0.37 | 29 (15.6) | 157 (84.4) | 0.17 | 28 (14.7) | 162 (85.3) | 0.32 |
| Three and more | 151 (41.7) | 56 (37.1) | 95 (62.9) |  | 30 (21.4) | 110 (78.6) |  | 26 (18.8) | 112 (81.2) |  |
| **Travel History** |  |  |  |  |  |  |  |  |  |  |
| Yes | 36 (9.9) | 13 (36.1) | 23 (63.9) | 0.63 | 7 (21.9) | 25 (78.1) | 0.55 | 10 (31.3) | 22 (68.8) | **0.01\*** |
| No | 326 (90.1) | 131 (40.2) | 195 (59.8) |  | 52 (17.7) | 242 (82.3) |  | 44 (14.9) | 252 (85.1) |  |
|  |  | Responder | Non-Responder | P-value | Responder | Non-Responder | P-value | Responder | Non-Responder | P-value |
| **Garbage pit within 200 metres** |  |  |  |  |  |  |  |  |  |  |
| Yes | 217 (59.9) | 99 (45.6) | 118 (54.4) | **0.005\*** | 37 (18.6) | 162 (81.4) | 0.77 | 27 (13.6) | 171 (86.4) | 0.08 |
| No | 145 (40.1) | 45 (31.0) | 100 (69.0) |  | 22 (17.3) | 105 (82.7) |  | 27 (20.8) | 103 (79.2) |  |
| **Vegetation within 200 metres** |  |  |  |  |  |  |  |  |  |  |
| Yes | 189 (52.2) | 91 (48.1) | 98 (51.9) | **<0.001\*** | 31 (18.6) | 136 (81.4) | 0.82 | 29 (17.3) | 139 (82.7) | 0.68 |
| No | 173 (47.8) | 53 (30.6) | 120 (69.4) |  | 28 (17.6) | 131 (82.4) |  | 25 (15.6) | 135 (84.4) |  |
| **Well/Stagnant water within 200 metres** |  |  |  |  |  |  |  |  |  |  |
| Yes | 153 (42.3) | 64 (41.8) | 89 (58.2) | 0.49 | 18 (13.7) | 113 (86.3) | 0.09 | 24 (17.5) | 113 (82.5) | 0.66 |
| No | 209 (57.7) | 80 (38.3) | 129 (61.7) |  | 41 (21.0) | 154 (79.0) |  | 30 (15.7) | 161 (84.3) |  |
| **SES** |  |  |  |  |  |  |  |  |  |  |
| Low | 290 (80.1) | 115 (39.7) | 175 (60.3) | 0.47 | 48 (18.3) | 215 (81.7) | 0.98 | 42 (15.9) | 222 (84.1) | 0.69 |
| Medium | 57 (15.7) | 25 (43.9) | 32 (56.1) |  | 9 (17.6) | 42 (82.4) |  | 9 (17.3) | 43 (82.7) |  |
| High | 15 (4.1) | 4 (26.7) | 11 (73.3) |  | 2 (6.7) | 10 (83.3) |  | 3 (25.0) | 9 (75.0) |  |
| **Housing Building Structure** |  |  |  |  |  |  |  |  |  |  |
| Mud | 210 (58.0) | 84 (40.0) | 126 (60.0) | 0.09 | 28 (14.8) | 161 (85.2) | 0.11 | 31 (16.3) | 159 (83.7) | 0.55 |
| Blocks | 90 (24.9) | 42 (46.7) | 48 (53.3) |  | 21 (25.3) | 62 (74.7) |  | 16 (19.5) | 66 (80.5) |  |
| Wood | 62 (17.1) | 18 (29.0) | 44 (71.0) |  | 10 (18.5) | 44 (81.5) |  | 7 (12.5) | 49 (87.5) |  |
| **Livestock more than 5** |  |  |  |  |  |  |  |  |  |  |
| Yes | 65 (18.0) | 29 (44.6) | 36 (55.4) | 0.37 | 15 (26.3) | 42 (73.7) | 0.07 | 15 (24.2) | 47 (75.8) | 0.06 |
| No | 297 (82.0) | 115 (38.7) | 182 (61.3) |  | 44 (16.4) | 225 (83.6) |  | 39 (14.7) | 227 (85.3) |  |
| **House Roofing** |  |  |  |  |  |  |  |  |  |  |
| Vigae | 56 (15.5) | 22 (39.3) | 34 (60.7) | 0.96 | 6 (13.3) | 39 (86.7) | 0.38 | 4 (8.5) | 43 (91.5) | 0.27 |
| Iron sheet | 269 (74.3) | 108 (40.1) | 161 (59.9) |  | 45 (18.0) | 205 (82.0) |  | 43 (17.6) | 201 (82.4) |  |
| Glasses | 37 (10.2) | 14 (37.8) | 23 (62.2) |  | 8 (25.8) | 23 (74.2) |  | 7 (18.9) | 30 (81.1) |  |

Rainy season (June 2021), Dry season (September 2021), (Short rainy (January 2022), SES: Social economic Status, \**p* computed by Chi-square test.

### Prevalence of Flavivirus and Alphavirus

Alphavirus positivity was highest during the rainy season, with 19 participants (5.2%) testing positive compared to 4 (1.1%) during the dry season and 9 (2.5%) during the short rainy season. For Flavivirus, positive cases were fewer overall. During the rainy season, 11 participants (3.0%) were positive, decreasing to 3 (0.8%) during the dry season, and 5 (1.4%) in the short rainy season, (Table 2).

**Table 2:** Prevalence of Alphavirus and Flavivirus infections by season in Northern Tanzania.

| **Virus** | **Rainy season**<br>**n (%)** |  | **Dry season**<br>**n (%)** |  | **Short rainy season**<br>**n (%)** |  |
| --- | --- | --- | --- | --- | --- | --- |
|  | **Positive** | **Negative** | **Positive** | **Negative** | **Positive** | **Negative** |
| *Alphavirus* | 19 (5.2) | 343 (94.8) | 4 (1.1) | 358 (98.9) | 9 (2.5) | 353 (97.5) |
| Flavivirus | 11 (3.0) | 352 (97.0) | 3 (0.8) | 359 (99.2) | 5 (1.4) | 357 (98.6) |

### Antibody Levels in PCR positive vs. PCR negative Individuals

For all three arboviruses investigated, the levels of IgG antibody responses against the Aedes 34 kDa Nterm peptide elevated in individuals testing positive for Flavivirus and Alphavirus compared to those testing negative (Mann-Whitney U test, *P* < 0.0001 for each comparison) (Figure 1).

**Figure 1.**
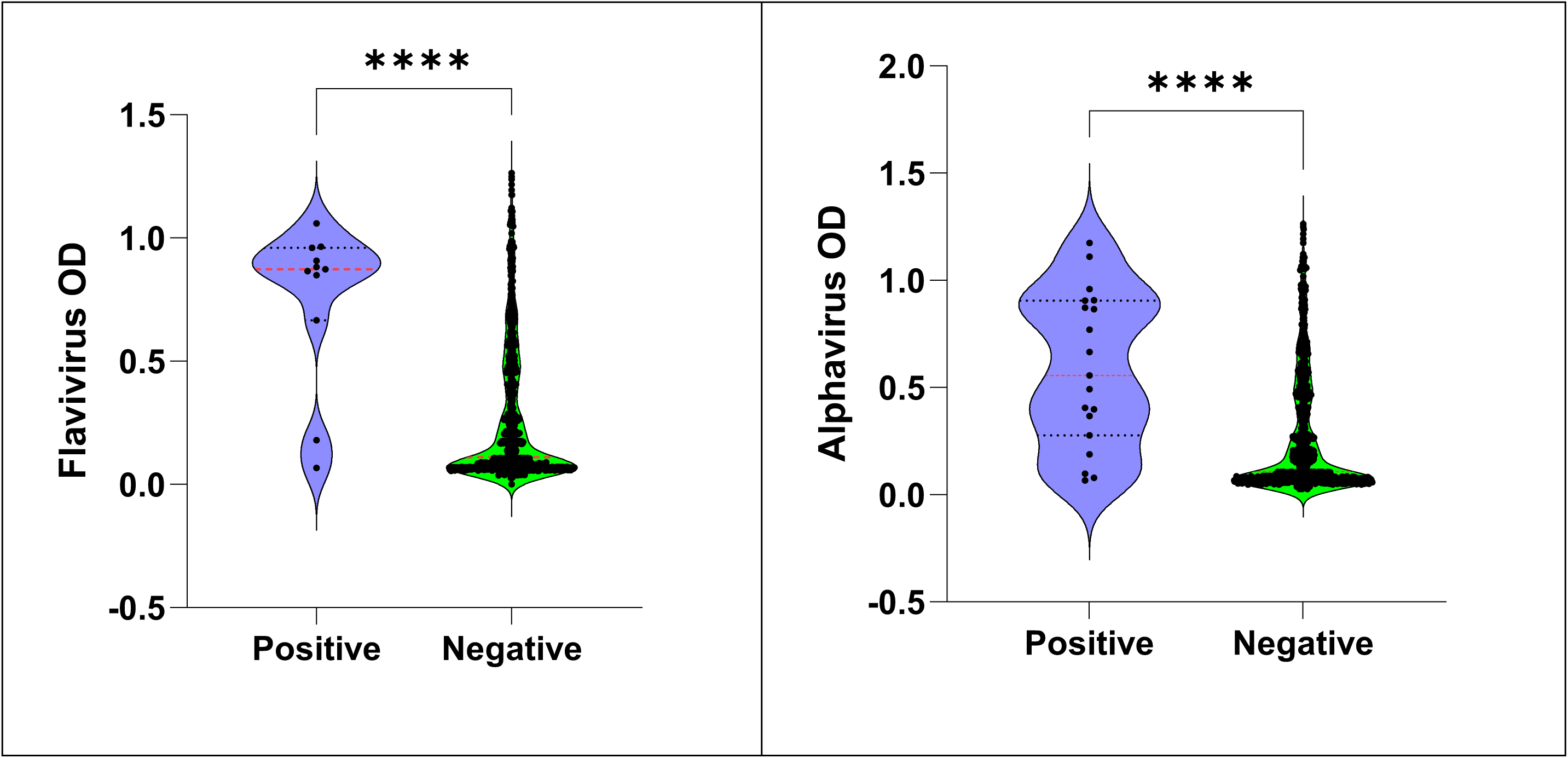
IgG antibody response to participants tested positive and negative by PCR against three arboviruses, dengue, zika and chikungunya in Tanga. Black points represent individual IgG responses (ΔOD), and red bars indicate the median value for each group. Statistical comparisons were performed using the Mann–Whitney U test. \*\**p* < 0.01; \*\*\*\**p* < 0.0001.

### Responders and non-responders to the 34kDa salivary peptide across different rainfall seasons

During the rainy season, among 362 participants, 144 (39.8%) were identified as responders, whereas 218 (60.2%) were classified as non-responders. In the short rainy season, of the 326 participants assessed, 59 (18.1%) were responders and 267 (81.9%) were non-responders. Additionally, among 328 participants evaluated during another short rainy season period, 54 (16.5%) were classified as responders, while 274 (83.5%) were categorized as non-responders (Figure 2).

**Figure 2:**
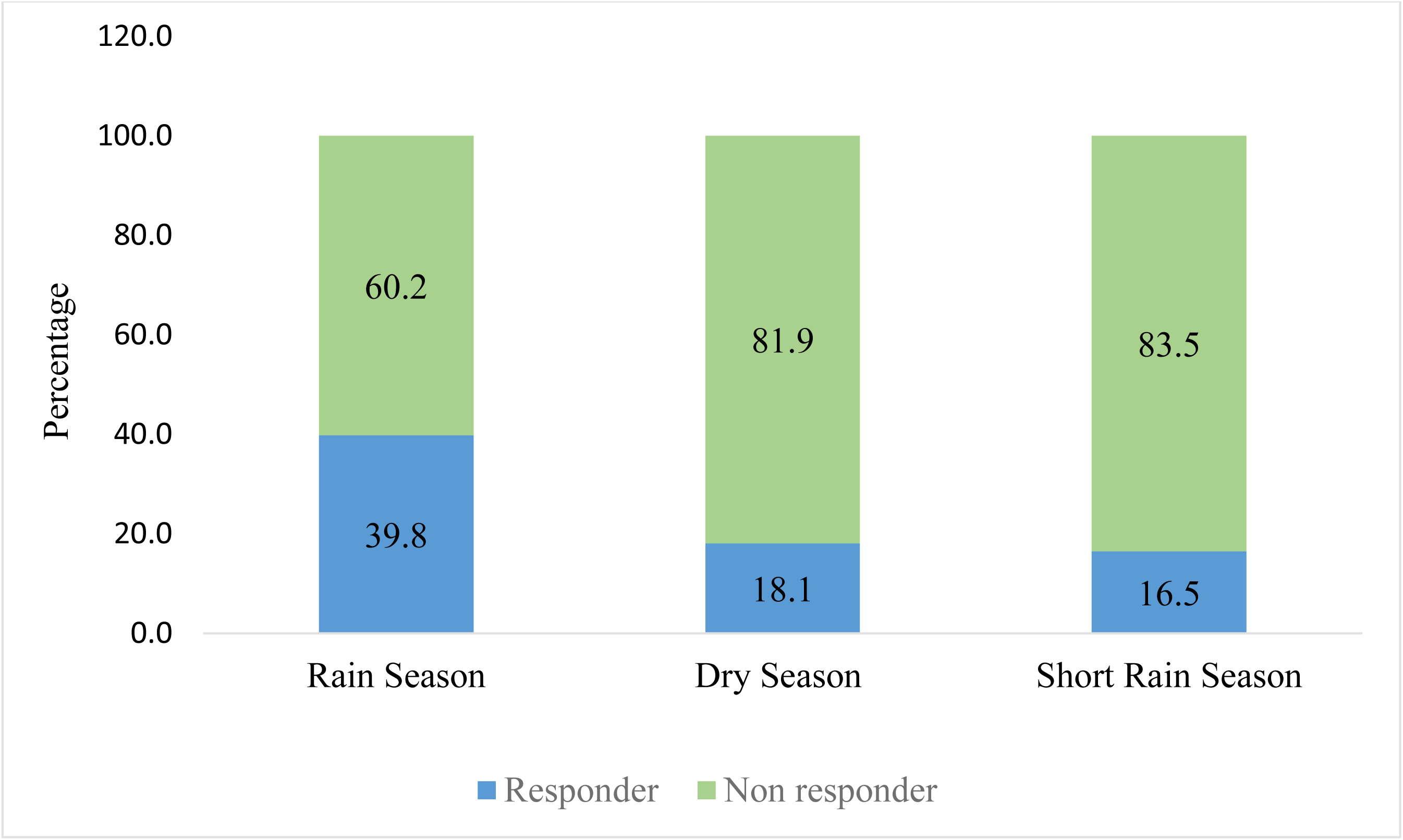
Distribution of IgG responders and non-responders to the 34kDa salivary peptide across different rainfall seasons in northern Tanzania.

### Seasonal Variation in *Aedes* NTerm 34 kDa IgG Antibody Levels

Mosquito bite exposure status was determined by classifying participants as responders or non-responders based on their salivary antigen antibody levels. A significant difference in median antibody levels was observed between seasons for those categorized as responders to the salivary biomarker. Specifically, individuals who responded to the antigen in the rainy and short rainy seasons demonstrated higher median antibody levels than those in the dry season (Mann-Whitney U test, *p* < 0.0001). Analysis of IgG antibody levels against the salivary biomarker *Aedes* Nterm 34 kDa revealed a highly significant difference across the rainy, dry, and short rainy seasons (Kruskal-Wallis test, *p* < 0.0001; Figure 3 and 4).

**Figure 3.**
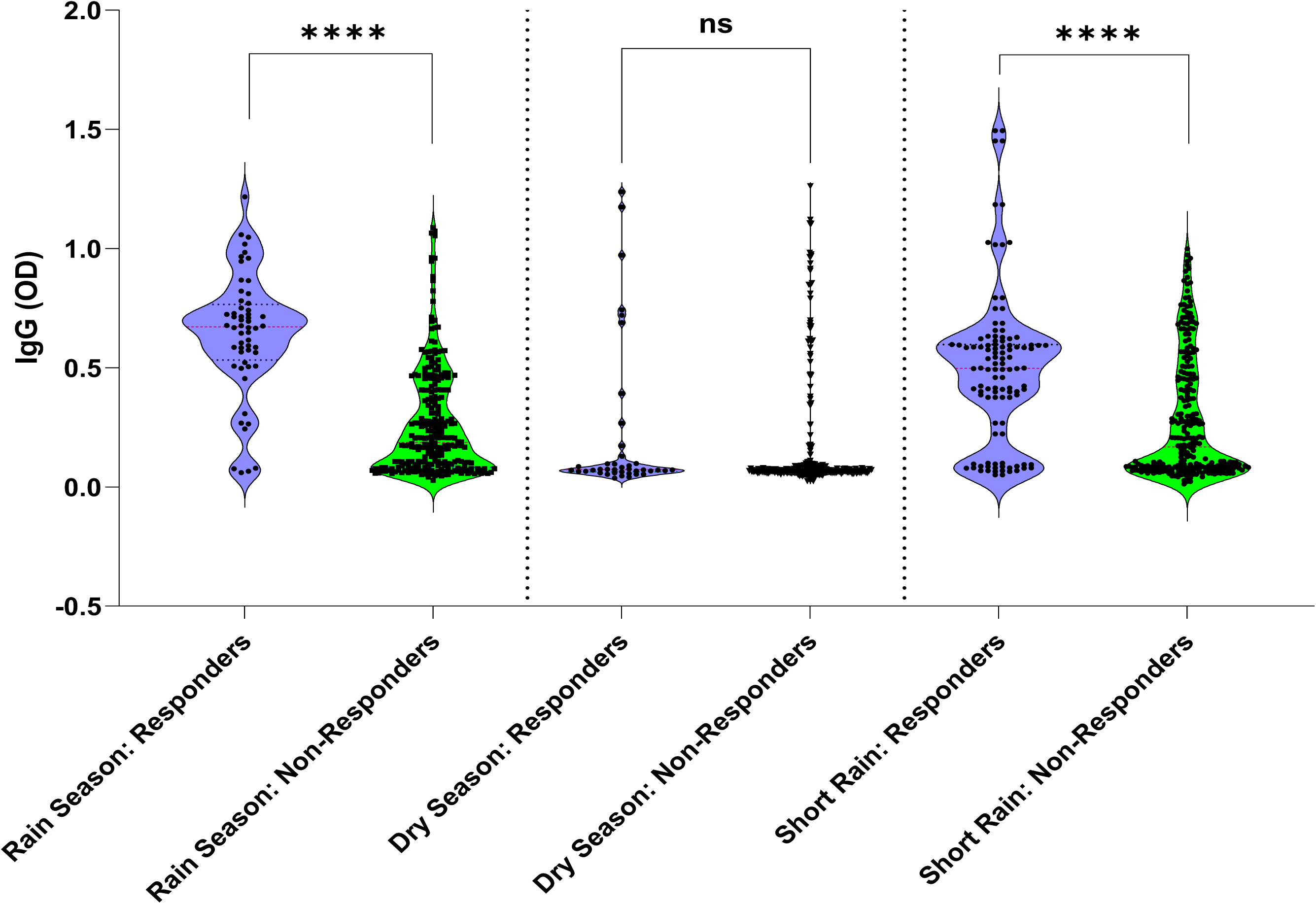
IgG antibody response to *Ae. aegypti* in responders and non-responders individuals across three seasons in Tanga. Black points represent individual IgG responses (ΔOD), and red bars indicate the median value for each group. Statistical comparisons were performed using the Mann–Whitney U test. \*\*\*\**p* < 0.0001, ns=not significant.

**Figure 4.**
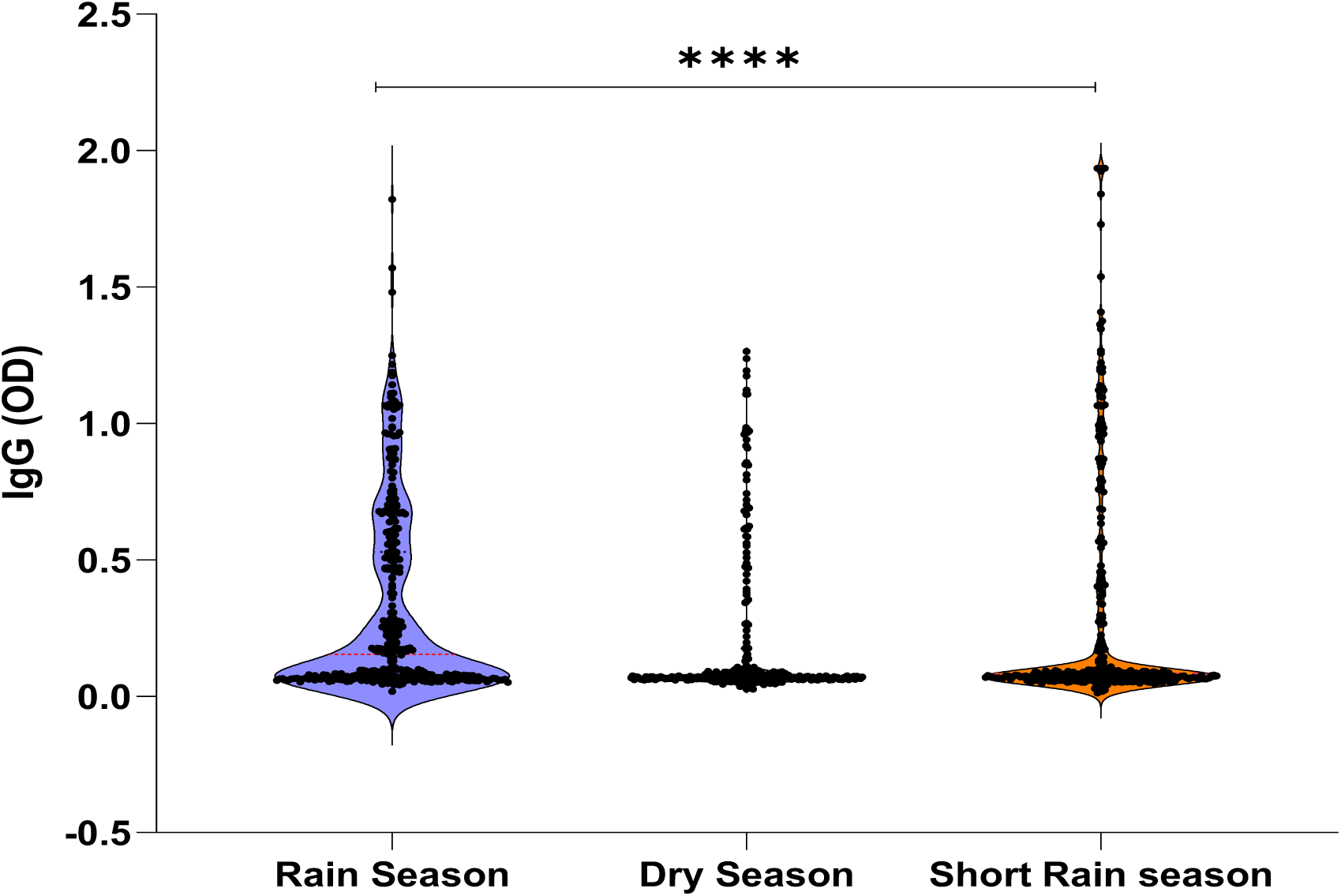
Individual IgG response to Nterm-34 kDa peptide between the rainy, dry and the Short rainy season. Results are presented during rainy season (June, 2021) the short rainy (October, 2021) and dry season (Feb 2022). Black points indicate individual IgG response (ΔOD) and red bars indicate the median value for each group and red bars indicate the median value for each group. Statistical comparisons were performed using the Kruskal-Wallis test. \*\*\*\**p* < 0.0001.

### Geographic and Seasonal Differences in *Aedes* Nterm 34 kDa IgG Antibody Levels

When assessing *Aedes* Nterm 34 kDa IgG antibody levels across three different villages and seasons, a statistically significant difference was observed specifically during the rainy season (Kruskal-Wallis test, P < 0.0001; Figure 5). However, no statistically significant differences were detected among villages during the dry and short rainy seasons.

**Figure 5.**
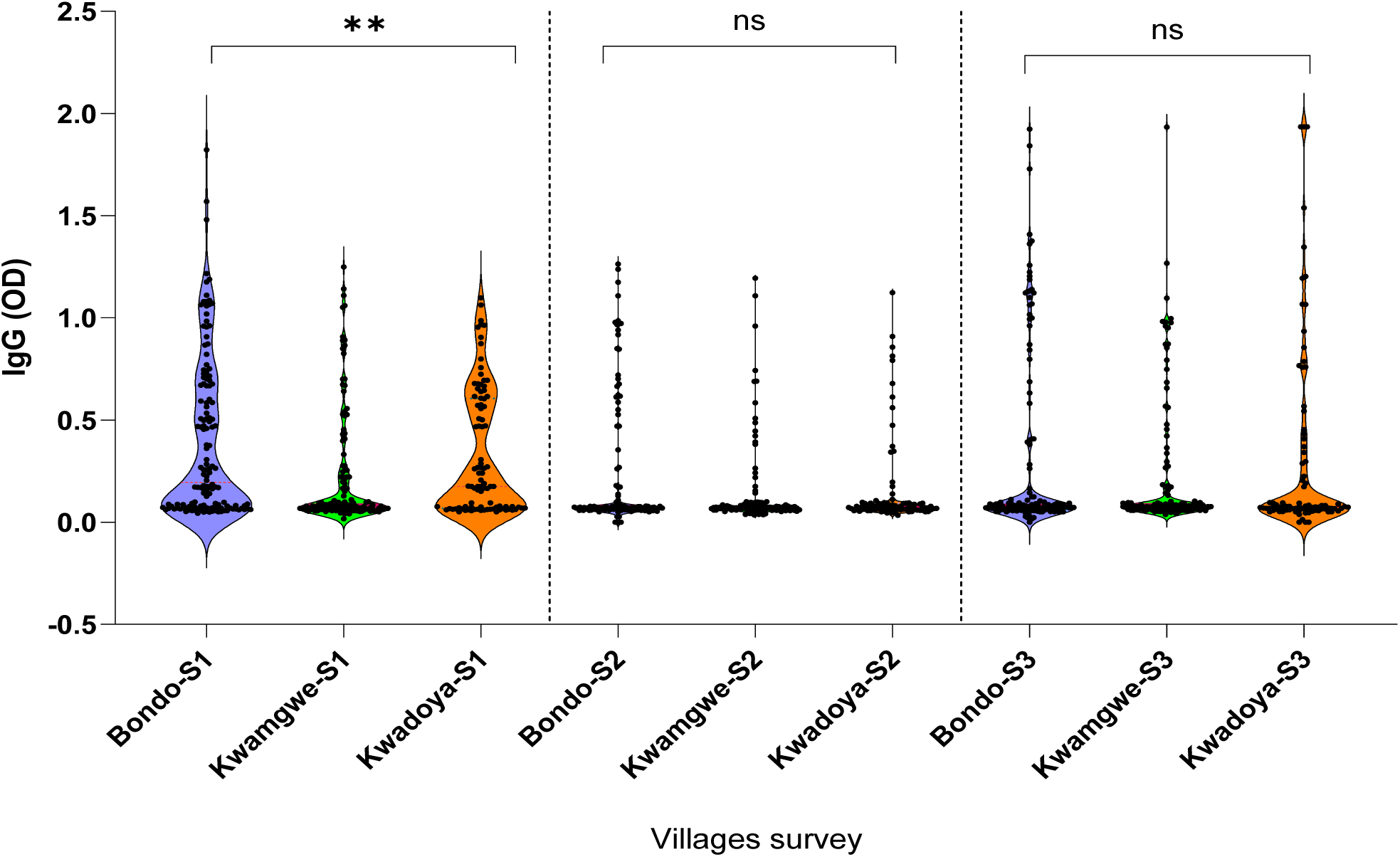
Individual IgG response to Nterm-34 kDa peptide between three villages across three seasons. Results are presented during rainy season (S1) (June, 2021) the short rainy (S2) (October, 2021) and dry season (S3) (Feb 2022). Black points indicate individual IgG response (ΔOD) and red bars indicate the median value for each group. Statistical comparisons were performed using the Kruskal-Wallis test. \*\*\*\**p* < 0.0001, ns=not significant.

### Factors associated with IgG seropositivity (Responders Non-responder)

During rainy seasons, participants residing near vegetation and garbage pits within 200 meters had higher seropositivity (*p* < 0.001 and *p* = 0.005, respectively). In short rainy, individuals with recent travel history showed significantly higher IgG responses (*p* = 0.01). Age, sex, income, occupation, and education level did not show statistically significant differences across surveys, though adults (≥18 years) consistently accounted for the majority of responders. Housing structure, livestock ownership, and roof type were not significantly associated with IgG seropositivity. These findings suggest a potential influence of localized environmental exposure and human movement patterns on short-term antibody responses (Table 1).

## Discussion

This study reports on the use of *Aedes* salivary biomarkers as a novel approach to assess the risk of arboviral diseases transmission. These biomarkers serve as proxies for mosquito exposure, enabling a more precise evaluation of infection risk within the population. *Aedes* salivary biomarkers offer potential as an early warning system by detecting mosquito exposure before clinical symptoms appear, aiding health authorities and policymakers in planning and implementing timely interventions. This study investigated antibody levels against the Nterm 34 kDa antigen to assess the risk of alphavirus and flavivirus, considering seasonal and geographic variations. The findings provide valuable insights into arbovirus transmission dynamics and demonstrate the utility of salivary antigens as indicators of human-vector contact, addressing the critical need for robust surveillance.

The results show that median antibody levels for all three arboviruses were significantly elevated in PCR-positive individuals compared to PCR-negative individuals. This finding confirms the expected antibody response following exposure to mosquito bites and infection with these viruses, reinforcing the reliability of salivary biomarkers in assessing the risk of arboviral infections. This is particularly relevant in the African region, where there are increasing reports of arboviral outbreaks. The same findings were reported In Columbia among dengue patients (Londono-Renteria et al., 2013; Cardenas et al., 2019; Elanga-ndille at al., 2013). The significant difference in median antibody levels observed between seasons among individuals responders to mosquito bites highlights the dynamic nature of human-vector interaction. Specifically, the higher median antibody levels during the rainy and short rainy seasons compared to the dry season strongly suggest increased mosquito biting rates during periods of higher precipitation. Further analysis of IgG antibody levels against the *Aedes* Nterm 34 kDa salivary biomarker revealed a highly significant seasonal variation across the rainy, dry, and short rainy seasons

This pattern provides robust evidence for fluctuations in vector activity throughout the year, with peak human exposure likely occurring during the rainy and short rainy seasons (Elanga-Ndille et al. 2012; Etienne et al. 2023; Fustec et al. 2021). These seasonal differences in vector exposure are critical for understanding the seasonality of arbovirus transmission and for informing public health interventions, such as targeted vector control campaigns during periods of high transmission risk, this is in line with a study done in Reunion Island which reported the importance of salivary biomarkers (Doucoure et al. 2012). Also, this finding aligns with the known ecology of *Aedes* mosquitoes, which typically thrive and exhibit higher population densities during rainy seasons due to increased breeding sites—a characteristic feature of the Tanga region’s climate (Kahamba et al. 2020).

When examining geographic and seasonal differences in *Aedes* Nterm 34 kDa IgG antibody levels across three surveyed villages Bondo, Kwamgwe, and Kwadoya. A statistically significant difference was observed with during the rainy season. This suggests that even within relatively close geographic proximity, there can be localized variations in mosquito biting intensity during peak vector activity periods. Such spatial heterogeneity could be influenced by local environmental conditions (e.g., availability of stagnant water, vegetation density), socio-economic factors, housing structures, and the effectiveness of local vector control efforts (Poinsignon et al. 2025b).The lower median antibody during dry and short rainy indicates a more uniform, perhaps lower, level of mosquito exposure across these locations when overall vector activity is reduced. This highlights the importance of localized surveillance and intervention strategies, rather than a "one-size-fits-all" approach, particularly during high-risk seasons.

Furthermore, environmental and human behavioral factors have emerged as significant predictors of IgG seropositivity against *Aedes* bites across the three survey rounds, underscoring the importance of localized risk exposures. In rainy season, participants residing within 200 meters of vegetation and garbage pits exhibited significantly higher seropositivity (p < 0.001 and p = 0.005, respectively), highlighting the role of micro-environmental conditions in sustaining *Aedes.*mosquito breeding habitats, reflecting findings from Zanzibar where solid waste in containers and vegetation were identified as principal predictors of *Ae. aegypti* abundance and arbovirus transmission risk (Kampango et al. 2021).

In short rainy, individuals with recent travel history showed significantly elevated IgG responses, indicating transient exposure in high-transmission settings. This aligns with evidence that travel can introduce individuals to areas with elevated vector density and insufficient control, leading to measurable increases in anti-*Aedes* antibody levels.

No significant associations were found between IgG seropositivity and housing structure, livestock ownership, or roof type. This suggests that localized environmental features and human movement patterns play a more dominant role in short-term antibody responses than structural home characteristics. *Aedes* vectors’ ubiquitous presence in informal waste and water container habitats further emphasizes this dynamic (Saleh et al. 2020).

Conversely, demographic characteristics including age, sex, income, occupation, and educational level were not significantly associated with IgG seropositivity in any of the survey rounds, despite adults (≥18 years) constituting the majority of responders. This pattern is consistent with findings several studies with the Nterm-34kDa peptide were no difference was observed in the level of the immune response for these parameters. This reinforces the notion that once environmental and occupational exposures are controlled for, basic demographic variables contribute minimally to arbovirus antibody levels.

## Conclusion

This study demonstrates distinct seasonal patterns in human IgG antibody responses to the Aedes Nterm-34kDa salivary peptide, correlating with the prevalence of Alphavirus and Flavivirus infections in northern Tanzania. The highest seropositivity and viral detection occurred during the rainy season, coinciding with increased vector exposure linked to environmental factors such as proximity to vegetation and garbage pits. Our findings highlight the utility of the Nterm-34kDa peptide as a sensitive biomarker for monitoring arboviral transmission dynamics. Despite the predominance of low socioeconomic status and limited mobility among participants, localized environmental conditions significantly influenced exposure risk. These results emphasize the need for integrated vector management tailored to seasonal and micro-environmental variations to reduce arboviral disease burden. This work provides critical insights for surveillance and targeted interventions aimed at mitigating the impact of Aedes-borne viruses in Tanzania and similar ecological settings.

## Data Availability

Due to ethical and privacy restrictions related to patient information, the data underlying the findings of this study are not publicly available. The data are available from the corresponding author upon reasonable request. Requests will be reviewed by the Kilimanjaro Christian Medical University College Ethics Committee to ensure compliance with privacy and consent regulations.

## Acknowledgement

We thank all participants for their readiness and for taking part in the study. We acknowledge the efforts of the staff of Bondo Dispensary and Kwamgwe Health Centre for their dedication to data collection. We also thank the community leaders of the Kwamgwe ward for making this study successful.

## Author Contributions

DCK contributed to the conceptualization of the study, secured funding, collected data, performed the experiments, and wrote the original draft of the paper. NK performed the experiments and contributed reagents, materials, and analytical tools. ETI analyzed the data and contributed to data visualization. BE reviewed and edited the paper and also contributed to writing. NAK performed the experiments and provided reagents, materials, and analysis tools. EEN contributed to the conceptualization of the study, secured funding, wrote the paper, and participated in the review and editing process.

